# Climate Risks to Water, Sanitation and Hygiene Services and Evidence of Inclusive and Effective Interventions in Low and Middle-Income Countries: A Scoping Review

**DOI:** 10.1101/2024.08.21.24312122

**Authors:** Jane Wilbur, Doug Ruuska, Shahpara Nawaz, Julian Natukunda

## Abstract

People with disabilities face significant disparities in access to water, sanitation, and hygiene (WASH) services, negatively affecting their health. Climate change exacerbates this by damaging WASH infrastructure and disrupting behaviours. Despite their heightened vulnerability, the needs of people with disabilities are often overlooked in climate change response strategies, putting them at greater risk.

This study explored how climate change impacts WASH services and behaviours and whether climate-resilient WASH interventions are disability-inclusive and gender-equitable in low—and middle-income countries.

Nine online databases were searched in July 2023 and May 2024 to identify peer-reviewed papers (CINAHL Complete, Embase, Global Health; Web of Science; ECONLIT; DESASTRES, GreenFILE, MEDLINE via PubMed, ERIC - Education Resources Information Centre). Grey literature was identified through OPENGrey, WHO, AHRQ, BASE and Google Scholar. Eligible papers included data on the impact of weather or climate on WASH services and behaviours, particularly for people with disabilities and women. Studies focused on community-based WASH interventions in LMICs and were published between 2000 and 2023 in English.

Twenty-two studies were included. Thirteen included results about women and girls; two considered disability. Only two evaluated climate-resilient WASH interventions (rainwater harvesting), and neither focused on disability. Most studies examined rainfall uncertainty and drought, covering diverse locations, including South Asia and East Africa. Most studies were published after 2020. Results show that climate change is exacerbating WASH inequalities, particularly affecting women and people with disabilities, while also adversely impacting public health by disrupting water availability, quality, and sanitation services. Variability in rainfall, droughts, floods, and saltwater intrusion significantly affect water reliability, quantity, and quality, leading to increased waterborne diseases, mental health issues, and other health problems. Social capital and kinship networks are critical during water scarcity. People with disabilities are especially vulnerable, often relying on people feeling morally obligated to support water collection. To cope, people diversify their water sources and prioritise water use for agriculture and livelihood security over hygiene, which elevates health risks. Extreme weather events further complicate the situation by damaging sanitation facilities, leading to increased open defecation and the spread of diseases. Rebuilding sanitation facilities is often deprioritised due to repeated damage, mental fatigue of constant reconstruction, and immediate survival needs. Water is frequently prioritised for agriculture over proper sanitation and hygiene practices, resulting in higher rates of open defecation and declining hygiene, as water use for handwashing, cleaning utensils, laundry, and menstrual hygiene is restricted. Consuming saline water is associated with high blood pressure, hypertension, pre-eclampsia, and respiratory infections. Women and girls who wash menstrual materials and bathe in saline water risk skin burns, rashes, blisters, and urinary tract infections. The role of governments and service providers in facilitating adaptation was inconsistent, with a lack of focus on community engagement and equitable service delivery.

Climate change disproportionately impacts vulnerable populations’ access to WASH services. This review highlights the urgent need for research on climate-resilient WASH interventions, especially those addressing the needs of people with disabilities. Targeted support, sustainable management, and robust evidence are essential to building resilience and equality.

## Introduction

Access to water, sanitation, and hygiene (WASH) and related positive behaviours are essential for maintaining health and overall well-being [1–3]. Handwashing with soap and water can reduce acute respiratory infections—the world’s leading cause of morbidity and mortality—by 17% [4]. Improving access to WASH reduces childhood mortality from any cause by 17% and diarrhoea deaths by 45% [5]. Sanitation interventions can enhance quality of life and prevent diseases. A study in unsewered, low-income areas of Maputo, Mozambique, found that people who used ‘high-quality’ shared latrines reported higher safety, privacy and mental health levels than users of ‘low-quality’ pit latrines [6]. High-quality latrines had pour-flush systems, concrete structures, and lockable doors, while low-quality ones had unlined pits and minimal privacy. Despite these physical and mental health benefits, nearly one in five people globally lack safe sanitation, and almost one in ten do not have access to safely managed water [7].

Inequalities within WASH access are marked. Individuals with disabilities often face limited or lower-quality access to WASH services, leading to disproportionate adverse health outcomes [8–11]. Despite often facing poorer access, many people with disabilities have a greater need for WASH services than those without disabilities. This includes individuals with health conditions like incontinence, epilepsy, albinism, and skin diseases, as well as those who use orthotics, prostheses, or other assistive devices that require cleaning [12–17].

Globally, 1.3 billion people with disabilities worldwide are particularly vulnerable to climate change effects [18, 19]. They are more likely to live in disaster-prone areas and are up to four times more likely to die in a natural disaster compared to those without disabilities [18, 19]. This is partly caused by barriers to participating fully in public life, which impacts their capacity to communicate their requirements and obtain crucial safety information during extreme weather and disasters [20, 21]. Despite these risks, the needs of people with disabilities are rarely considered in climate responses. A systematic review of climate adaptation responses found that only 1% of the 1,680 articles reviewed considered people with disabilities; the least attention given to all marginalised groups assessed [22]. Furthermore, 81% of State parties to the Paris Agreement did not mention disability in their national plans, thus risking perpetuating existing inequalities [23].

The threats of climate change to WASH services and behaviours are increasingly evident. Rising sea levels, extreme heat, altered precipitation patterns, flooding, and cyclones disrupt water availability, quality, and demand. Surface water sources become less predictable while groundwater demand surges. Flooded sanitation facilities contaminate water and the environment, damaging WASH infrastructure like water points, latrines, and sewerage systems [6, 24–26]. These disruptions directly impact WASH behaviours: people may resort to coping mechanisms that include open defecation, unsafe water consumption, and long, often difficult journeys to collect water, especially in extreme heat. Menstrual health, handwashing, and personal hygiene practices decline, increasing health risks [27, 28]. People with disabilities, already facing significant barriers to WASH services, become even more vulnerable to the adverse health effects of climate change-induced WASH disruptions, potentially leading to severe health and social consequences.

This scoping review aimed to explore how climate change impacts WASH services and behaviours and if climate-resilient WASH interventions are disability-inclusive and gender-equitable in low and middle-income countries (LMICs). The research questions were: 1) What is the evidence that climate risks (including weather events) affect women and men with disabilities’ physical and mental health and well-being through disruption of WASH? and 2) What is the evidence for the effectiveness of climate-resilient WASH interventions in LMICS?

WASH interventions are defined as individual, household or community activities that promote health through effective hygiene behaviours. As set out in the UN Human Rights to Water and Sanitation, interventions should improve the availability of water and sanitation facilities, quality, acceptability, accessibility and affordability [29]. Positive sanitation behaviours include building a latrine and stopping open defecation. Effective hygiene behaviours involve hand hygiene, menstrual health, food hygiene, personal care and safe water collection and storage practices that help maintain health and prevent disease spread. Persons with disabilities are ‘those who have long-term physical, mental, intellectual or sensory impairments which in interaction with various barriers may hinder their full and effective participation in society on an equal basis with others’ [30]

Climate-resilient WASH is less clearly defined. We include any WASH programming that helps ensure that WASH infrastructure, services and behaviours are sustainable and resilient to human-induced climate change, including more intense and frequent weather events [31].

Achieving climate-resilient WASH necessitates a comprehensive, collaborative approach involving all stakeholders. This includes marginalised groups such as persons with disabilities, women, and girls, as well as households, communities, government entities at all levels, service providers, and regulators. By working together, these stakeholders can effectively identify, assess, and mitigate the risks posed by climate-related events to WASH services and the broader systems that support them. By fostering collaboration, climate-resilient WASH not only safeguards essential services but also plays a pivotal role in strengthening overall community resilience to the impacts of climate change [32].

## Materials and Methods

Scoping reviews are conducted when evidence has not yet been reviewed or is heterogeneous. They aim to identify and map the existing evidence on a topic of interest, identify and analyse knowledge gaps, and proceed with a systematic review [33–35]. This scoping review follows Peters, M. et al.’s guidance for conducting systematic scoping reviews [35]. A review protocol was registered online with OSF Registries (https://doi.org/10.17605/OSF.IO/SZCGB) and is reported in line with the PRISMA Extension for scoping reviews (S1, PRISMA checklist).

### Eligibility criteria

The search strategy was designed to identify peer-reviewed and grey literature of primary research that explored climate risks to WASH for people with disabilities in low- and middle-income countries. Eligible participants were women, girls, men and boys with and without disabilities. Studies were included if they met the following criteria:

1. Peer-reviewed and grey literature, including observational studies, case studies and reports that include primary data and reviews.
2. Studies that reported weather or climate impacts on WASH services and behaviours, including those disaggregated by disability and/or gender.
3. Papers that assessed the effectiveness of climate-resilient WASH interventions in community settings, defined as WASH services shared by households (e.g. shared handpumps/latrines) and within individual households.
4. No restrictions were placed on study design and included rapid reviews, process and outcome evaluations, feasibility studies and case studies or reports.
5. Studies undertaken in LMICs as defined by the World Bank country classification [36].
6. Papers published between 2000 and 2023 and written in English.

Studies were excluded if they:

- Did not address WASH services shared by households (such as urban systems or watershed studies).
- Focused on WASH services for people in temporary accommodation following a disaster, including displacement camps
- Reported flood events unrelated to the weather, such as dam bursts or tsunamis

#### Information sources

Nine online databases were searched: *CINAHL Complete, Embase*, Global Health; Web of Science; *ECONLIT; DESASTRES*, GreenFILE, MEDLINE via PubMed, and ERIC (Education Resources Information Centre). Grey literature was searched using OPENGrey, WHO, AHRQ, BASE and Google Scholar. Comprehensive search terms were created to capture four main concepts: weather or climate events, WASH, effectiveness, and disability (S2 Search strategy and key terms). Final search strings were peer-reviewed by academics experienced in the topics and the London School of Hygiene & Tropical Medicine (LSHTM) librarians.

#### Study selection

The search was conducted in July 2023 and rerun in May 2024. Different search strategy formulas were finalised to answer the two research questions. The first research question formula was: (Weather events terms) AND (WASH terms) AND (Evaluation terms) AND (time restriction 2000 to 2023) AND (restriction by English language). The second was: (Weather events terms) AND (WASH terms) AND (Disability) AND (time restriction 2000 to 2023) AND (restriction by English language). When an included article summarised findings from multiple primary studies, we manually searched for individual publications. Results were exported to EndNote X9 for de-duplication and then imported to Rayyan [37] for screening. One reviewer screened the titles, while the abstract and full-text screening process was blinded and conducted independently by all three reviewers. Any disagreements were discussed and agreed upon by all reviewers.

#### Data extraction and analysis

The following data were extracted into Microsoft Excel for analysis: country, aims/purpose, study population and sample size, methods, intervention type, concept, duration of the intervention, how outcomes were measured, and key findings related to the review questions. Due to the heterogeneous nature of the evidence gathered, applying a standardised reference scale to assess quality was impossible. This approach corresponds with guidance for conducting systematic scoping reviews, which states that, unlike systematic reviews, scoping reviews rarely formally evaluate the methodological quality of included papers [34, 35].

## Results

### Study selection

Twenty-two studies met our inclusion criteria (Figure 1 PRISMA flowchart). 1265 records were identified through electronic searches. After removing duplicates (n=333), 824 records were excluded during the title and abstract screen, leaving 108 studies for the full-text screen. Following a full-text review, 86 studies were excluded, and 22 were selected for inclusion.

**Fig 1:**
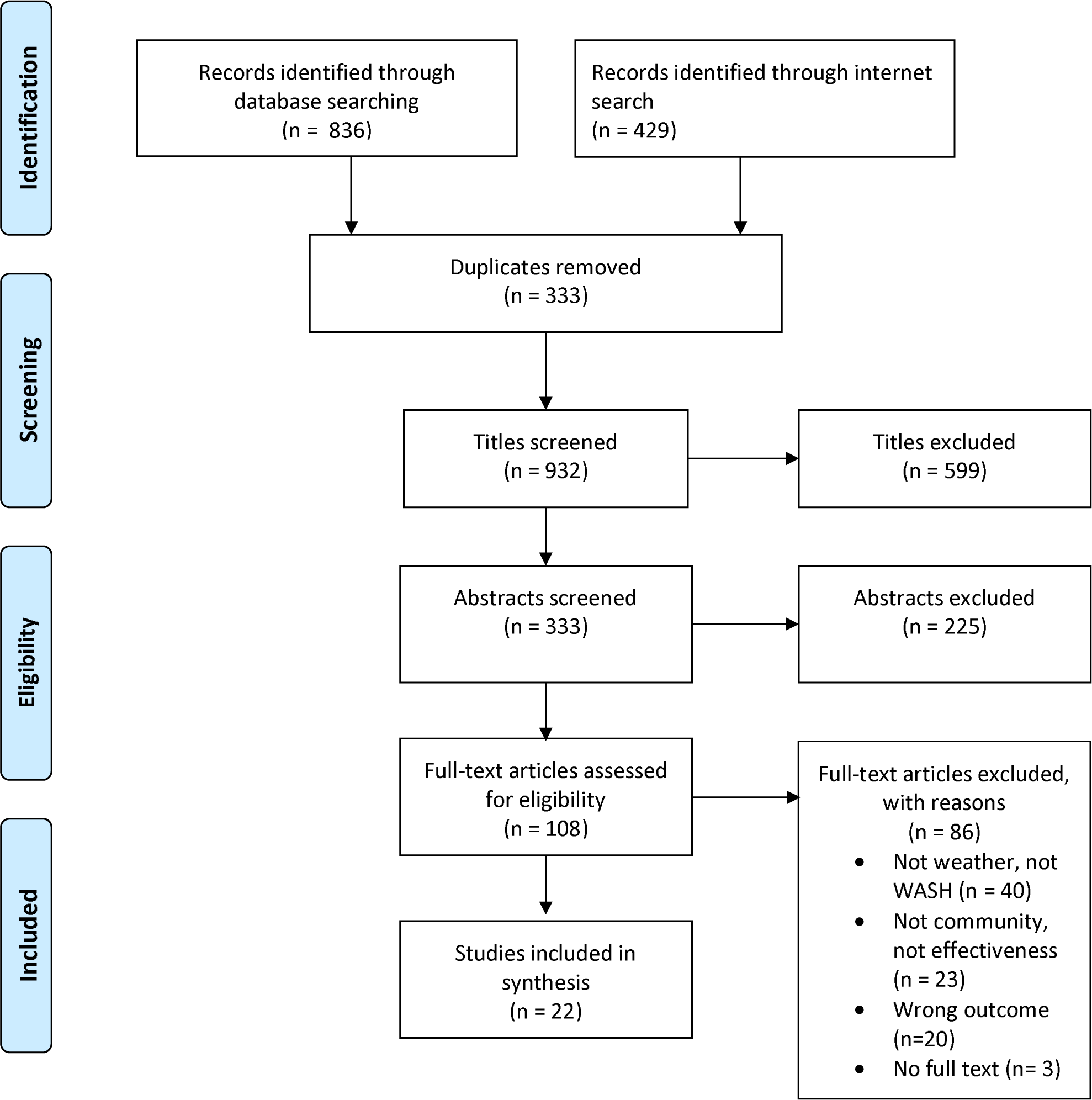
Search strategy with PRISMA flow diagram.

### Study characteristics

We did not find any studies that evaluated disability-inclusive climate-resilient WASH interventions, and there were minimal evaluations of climate-resilient WASH at the community or household level (n=2). There was more evidence regarding the health impacts and lived experiences of persons dealing with climate-driven water insecurity or damage to WASH infrastructure from weather and climate extremes (n=20). The types of climate hazards considered were predominantly rainfall uncertainty and drought (n=19). Papers were from a wide range of countries, with South Asia and East Africa most represented regions. There was an even distribution between rural and urban settings. Most papers (n=18) were published between 2020 and 2023.

### Weather events and impacts on health, WASH services and behaviours

Most papers documented low rainfall and drought, followed by heavy rain and flooding. Most studies addressed the impact of weather events on water services alone (n=6), and then equal attention was given to water and sanitation (n=5), water and hygiene (n=5), and water, sanitation, and hygiene (n=5). The fewest papers considered sanitation and hygiene together (n=1). Of the 22 papers, 13 included results about women and girls, and only two considered persons with disabilities.

Table 1 summarises studies that report how climate hazards impact population health and the coping strategies applied by the general population, women, girls, and people with disabilities. Coping strategies do not necessarily result in positive health outcomes. For instance, a coping strategy applied during water scarcity could be reducing handwashing, which could lead to increased infectious diseases.

**Table 1.**
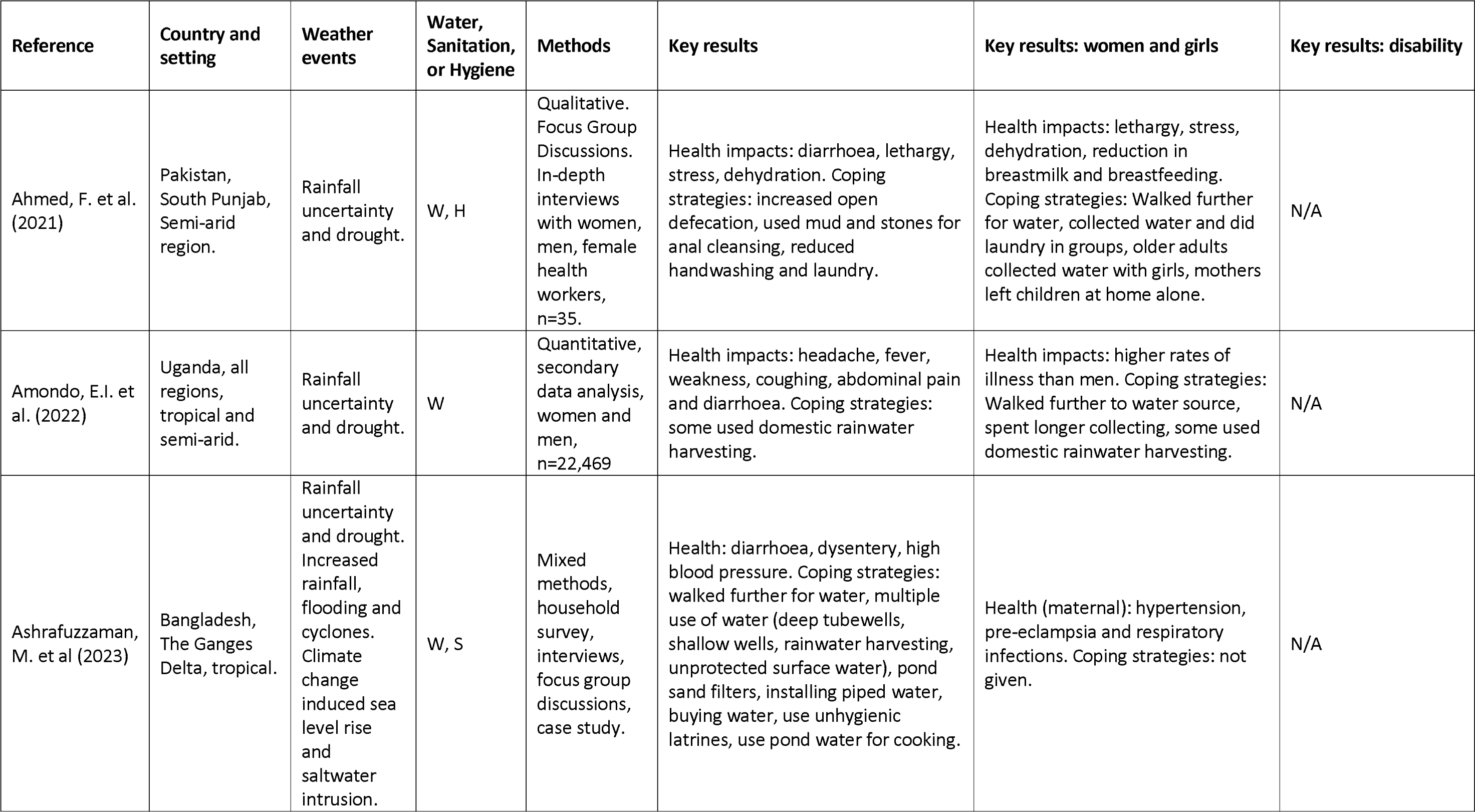
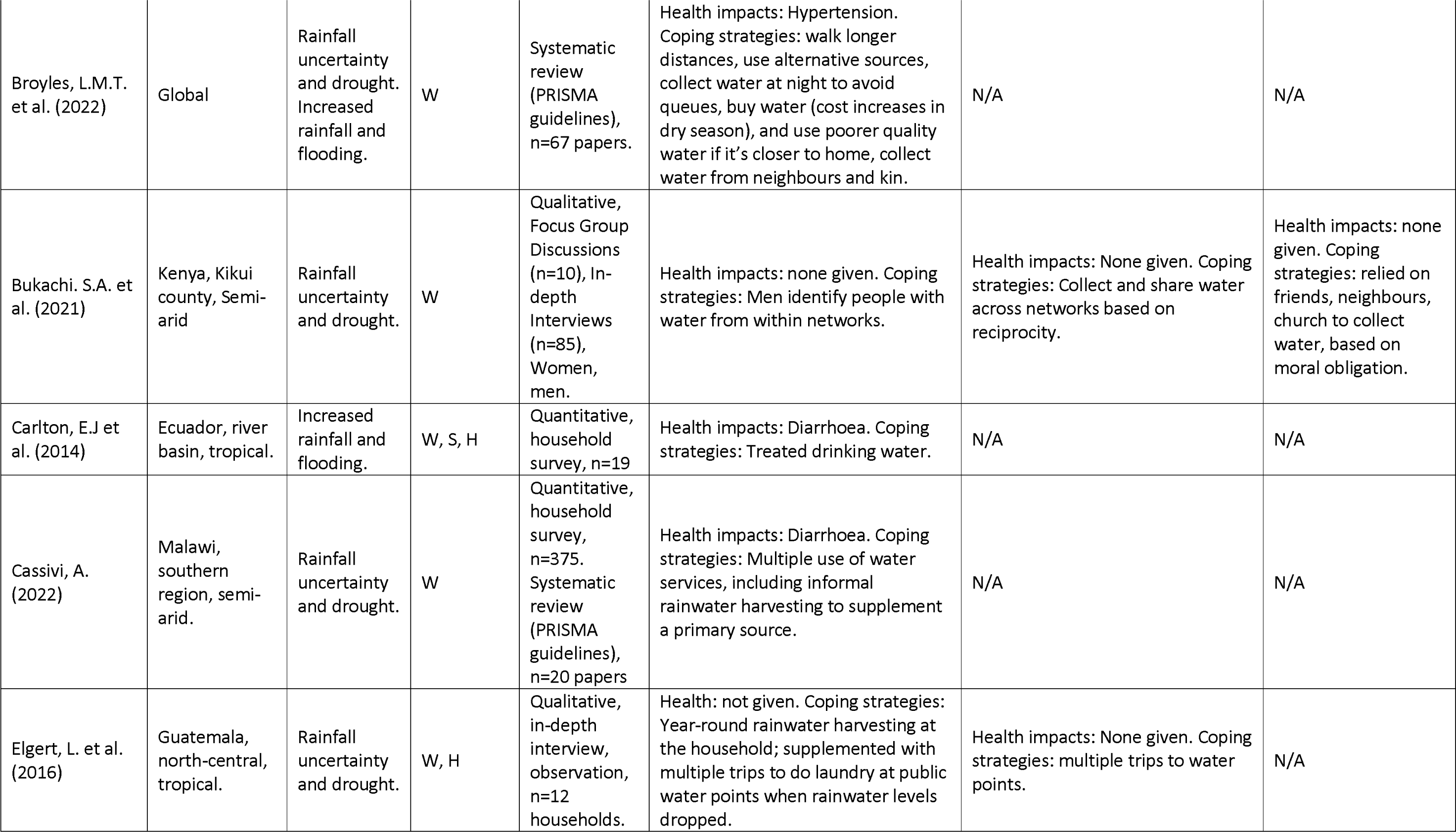
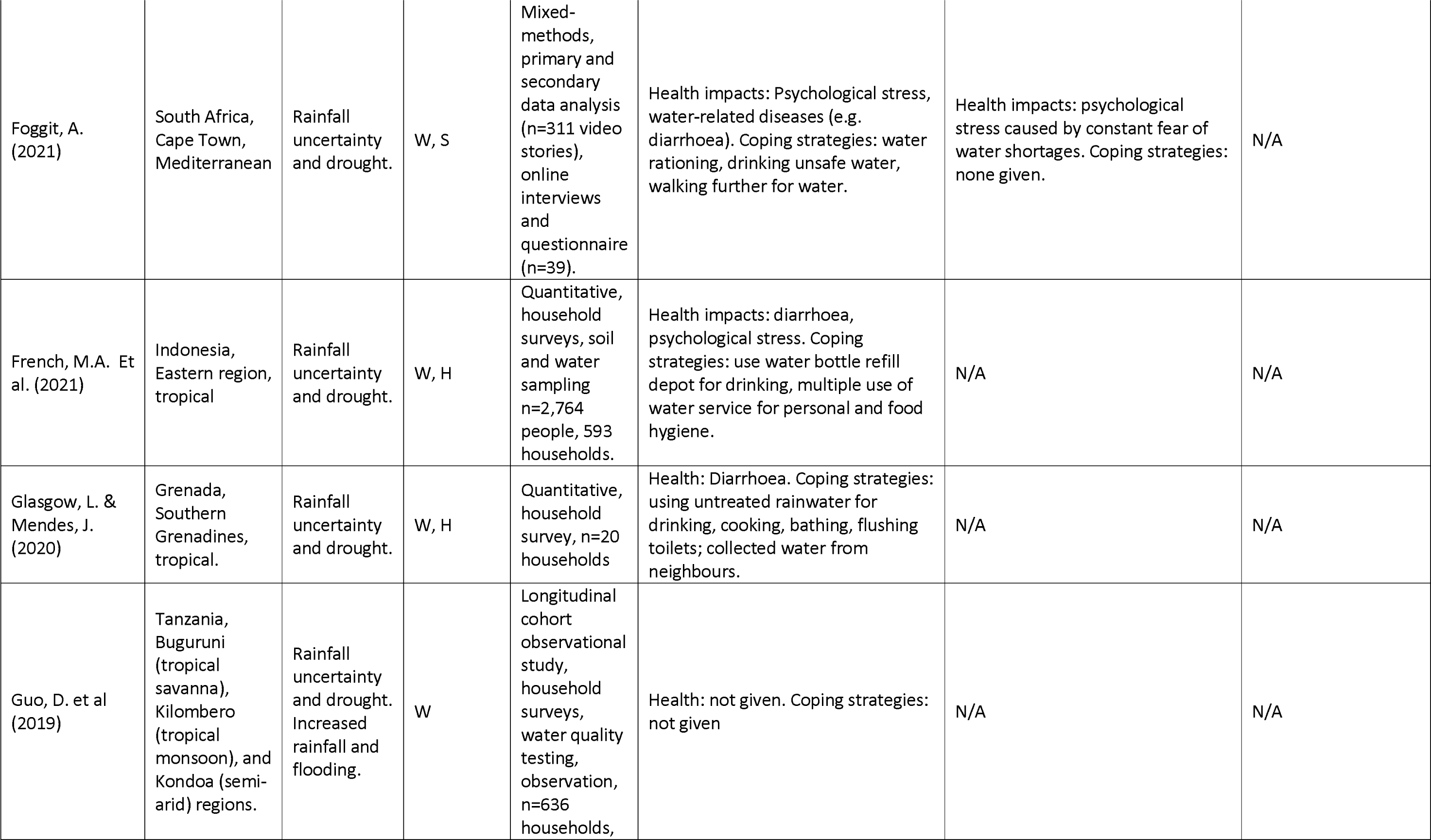
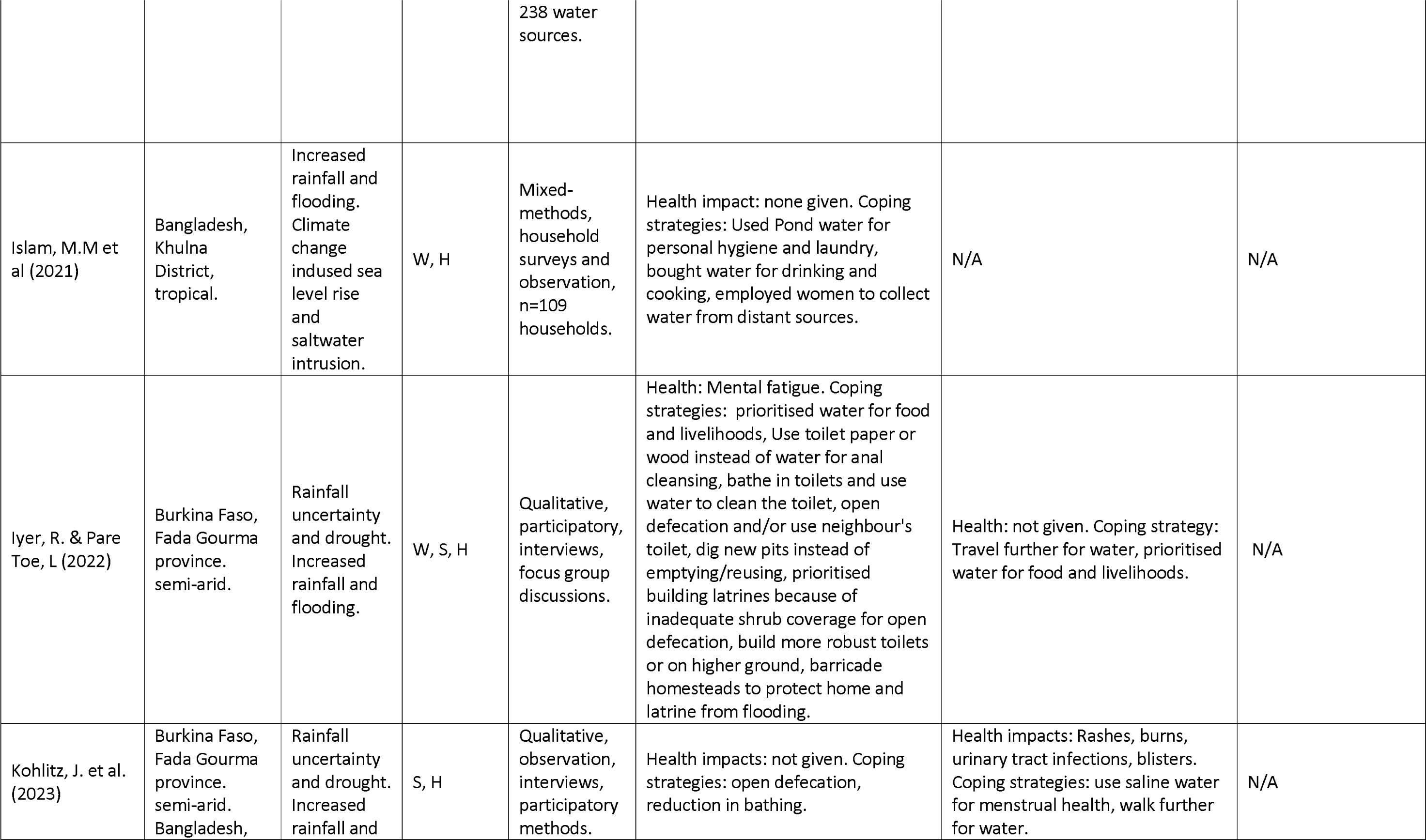
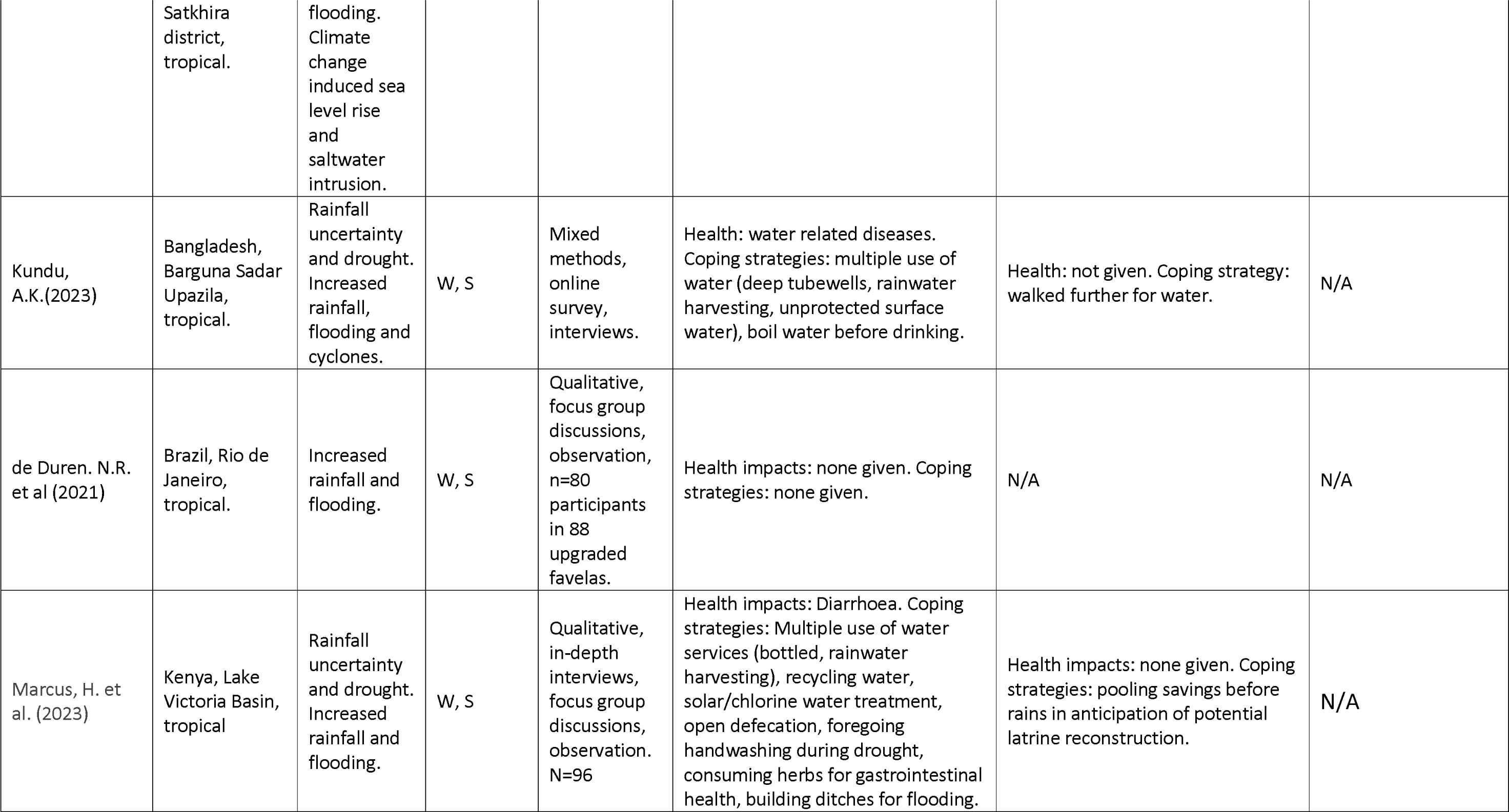
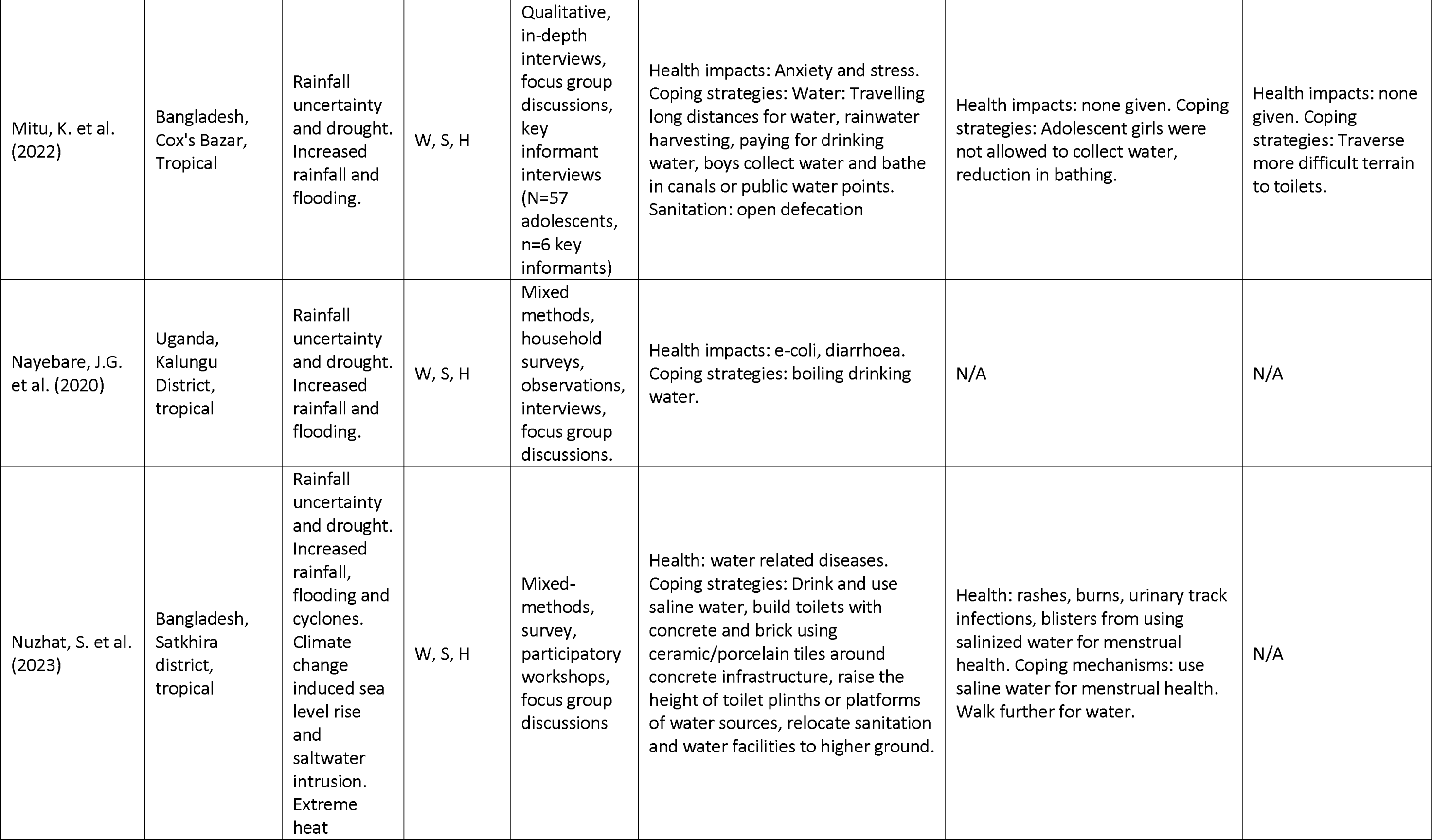
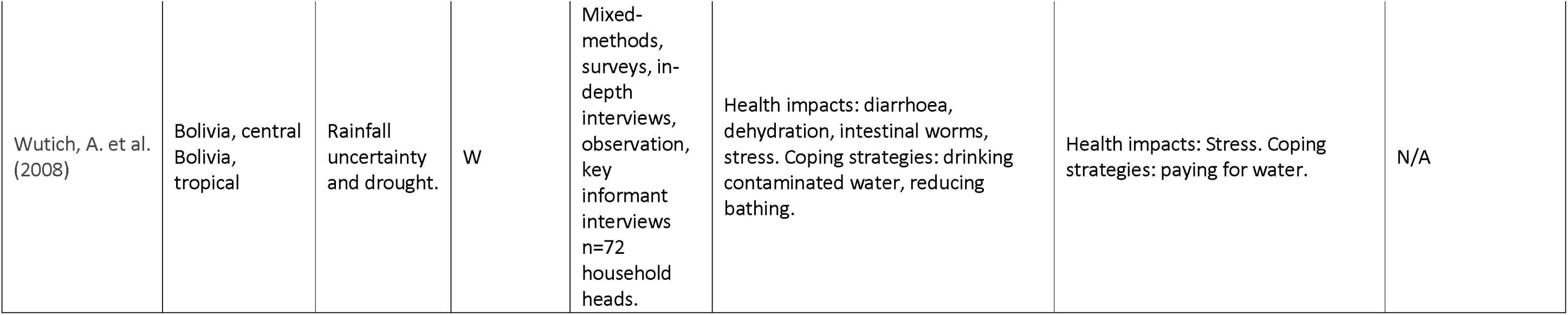
Summary of studies on impacts of climate hazards on WASH.

#### Rainfall uncertainty and drought

Papers reported that rainfall uncertainty and drought affect water availability’s reliability, quantity, and quality [38–47]. People responded by diversifying their water sources, including collecting water from rivers, rainwater, shallow wells, and vendors [46–48]. Some people requested water loans and borrowed or begged for water from neighbours [46, 48].

Bukachi et al. [49] highlighted the importance of social capital and kinship in facilitating water-sharing, subsidisation, or reciprocity between families, with gender dictating roles. Men identified a water source for the family, utilising their networks, friendships, neighbours and family relations, and women collected it and, or worked in exchange for water. People with disabilities who could not independently collect enough water relied on the moral obligation within social networks of friends, neighbours and the church for their water. People with disabilities were not expected to provide a service in return.

Drinking untreated rain and surface water was reported [46, 48]. Still, papers predominantly highlighted that households prioritised water for food and livelihood security above personal and food hygiene, such as handwashing, doing laundry, cleaning plates and baby feeding instruments [40, 44, 48, 50]. Studies reported a shift from using water for anal cleansing to paper, wood, mud and pebbles [40, 51].

Studies focusing on rainfall uncertainty and drought reported health impacts, including diarrhoea, abdominal pain, headache, dehydration, intestinal worms, and psychological stress [41, 45–48, 52]. Waterborne diseases, dehydration, and headaches were reported and linked to consuming contaminated water [45, 48, 52]. Declining mental health due to rainfall uncertainty and drought was reported in several papers. For instance, in Bolivia, household heads experienced worry, anger and fear because of water insecurity, which led to arguments [48]. Regarding gender roles, women’s burden increased during water shortages as they had to manage available water for cooking, cleaning, laundry, and the kitchen garden [51]. Women were particularly affected by the constant fear of water shortages [45]. One paper reported that men and women travelled further to collect water [38]. Still, most reported this role was carried out by women and girls who spent longer collecting water, covering greater distances, spending time in longer queues and making multiple trips [40, 52–54]. Women and girls faced harassment when collecting water, perpetrated by men, boys and other women queuing for water [40, 53, 54]. In South Punjab, Pakistan, water insecurity increased women’s and girls’ vulnerability to harassment and gender and caste-based violence, including sextortion [40]. Women were more stressed and lethargic, especially when sick or menstruating; dehydration was common, and lactating mothers were less able to produce milk in sufficient quantities, shortening breastfeeding [40].

#### Increased rainfall, flooding and cyclones

Heavy rainfall events and flooding led to wells collapsing [42–44, 55] and carrying built-up bacteria and other harmful organisms (pathogens) directly into surface water used for drinking and bathing [43, 56, 57]. This was particularly concentrated when dry periods, during which pathogens had built up in the environment, were followed by heavy rainfall, resulting in a significantly increased risk of E. coli in water and diarrhoea transmission [43, 57].

Some people treat drinking water by filtering and boiling it, using solar water disinfection, and adding chlorine [43, 44, 54, 57]. Other coping strategies include paying for drinking water and storing and recycling water. The most common approach was constructing rainwater harvesting systems at homes, used for drinking (if treated), bathing, laundry and cooking [44, 58].

Six papers reported that heavy rains, flooding, and cyclones damaged latrine infrastructure, waterlogging and overflowing latrines and drainage systems [38, 39, 44, 51, 54, 58]. For instance, Marcus highlighted how heavy rains lead to pit latrine damage and collapse in Kenya’s Lake Victoria Basin [44]. In Rio de Janeiro, Libertun de Duren et al. found that sewerage systems overflowed during heavy rains, but this was also caused by population growth overtaking the capacity of the sanitation system [59]. In Cox’s Bazar, Bangladesh, Mitu et al. reported that heavy rains, flash floods, and landslides caused waterlogging as drainage systems were not regularly cleared [58]. The authors also noted that adolescents with disabilities living in long-term Rohingya refugee settlements faced additional challenges reaching latrines because heavy rains damaged roads and paths [58]. Nuzhat et al. noted that women did not want to use damaged latrines that were dark and lacked privacy [39].

Many latrines were often damaged from previous weather events, so they could not withstand the latest incident [39, 51]. Sanitation was deprioritised for several reasons: 1) the recurring need to rebuild damaged latrines, causing mental fatigue and related costs after a weather event; 2) the inability to reach the local market to buy materials to rebuild the latrine because the roads are damaged by floods, 3) a focus on rebuilding the homestead or food and livelihood security [39, 51]. Studies reported continued use of unhygienic latrines [38, 39], increased rates of open defecation [39], using neighbours’ latrines, or a combination of the latter two options [51]. Due to impacts on sanitation services and behaviours, increased waterborne diseases, including diarrhoea and skin conditions, were reported [39, 54].

Some adaptation activities were documented [39, 44, 51]. People in Kenya built ditches and dams around houses for flood control, saved for latrine reconstruction ahead of the rainy season, constructed latrines in higher ground to increase resistance to flooding, and planted trees in household yards to improve soil water retention [44]. In Burkina Faso, some people built storm and flood defences around their homesteads for protection [51]. Following damage, those who could afford to build stronger, more flood-proof latrines used more durable materials such as cement, strong pit liners, and concrete slabs; some relocated latrines to higher ground or raised the height of the latrine plinths above flood levels [39, 44, 51].

Nuzhat et al. [39] included information about how heavy rainfall, flooding and cyclones impacted the ability of women and girls to manage menstruation. Damage to latrines affected people’s ability to change their menstrual materials in private, and heavy rains and flooding made it difficult to clean and dry reusable materials. Many people evacuated their homes without menstrual materials. They could not buy replacements as shops were closed during the weather event, and cyclone shelters lacked adequate water, sanitation and hygiene services to facilitate menstrual health.

#### Climate change-induced sea level rise and saltwater intrusion

Climate change-induced sea level rise and saltwater intrusion in freshwater was documented in three papers [38, 39, 42]. One study reported how high salinity affected sanitation service durability [39]. Households that could afford it constructed latrines’ infrastructure with concrete and installed ceramic or porcelain tiles on top [39].

All papers documented saline intrusion in groundwater, shallow wells and surface water in subtropical and tropical floodplain rivers, coastal rivers, and oceanic islands [38, 39, 42]. In Bangladesh, this reduced water for drinking, cooking and agriculture all year round [38, 39]. Regarding water, coping mechanisms applied included using multiple water sources, including deep tubewells, handpumps, shallow wells, rainwater harvesting, and surface water (rivers and ponds). Most households used rainwater to drink for two to three months a year and then shifted to salinated pond water [38]. The pond water was primarily used for cooking; some used community-level pond sand filtration. Those who could afford it bought water from Reverse Osmosis plants or installed piped water in their homes [38, 39, 60]. However, many people had to consume highly saline water [38, 39].

Health impacts of consuming highly saline water reported were high blood pressure, increased rates of hypertension, pre-eclampsia and respiratory infections [38]. Women and girls who bathed in and washed reusable menstrual materials in water with high saline content reported burns, rashes, blisters, and urinary tract infections [39]. Finally, high salinity in Bangladesh has changed livelihood practices. Previously, crop cultivation was common, but this is no longer a viable option because saltwater intrusion reduced soil fertility. Now, many people farm shrimp, but this further increases groundwater salinity [39]. Furthermore, many farmers cannot afford modern irrigation technologies, so they consume significant volumes of ground and surface water.

### Effectiveness of climate-resilient WASH at the household and community levels

There is minimal evidence regarding the effectiveness of climate-resilient WASH services at the household and community levels. Only two papers explored this and focused on rainwater harvesting technology (Table 2). No papers explored efforts to maintain positive WASH behaviours (e.g. safe water management, handwashing with soap and water at critical times, faecal management and disposal, food hygiene and menstrual hygiene) before, during or after a climatic event.

**Table 2.** Studies on the effectiveness of climate-resilient WASH.

| Study | Study aim | Study site and methodology | Technology | Key findings |
| --- | --- | --- | --- | --- |
| Elgert, L. et al. (2016) | Evaluate if rainwater harvesting has improved water security in a peri-urban village in Guatemala and draws implications from this project for expansion to the region and beyond. | Peri-urban village in Guatemala. Qualitative: interviews and observation of rainwater harvesting technology, 12 households. | Household rainwater harvesting | Increased household water security and a reduction in time and energy expended by women collecting water from public water points. However, some negative impacts regarding cost, water quality. |
| Glasgow, L. & Mendes, J. (2020) | Assess the knowledge, attitudes and practices about household rainwater harvesting. | Carricoao, Grenada. Quantitative: survey of 20 households in 15 communities. | Household rainwater harvesting | Increased water security but some concerns about water quality associated with poor maintenance. |

Elgert et al. [53] explored whether household rainwater harvesting improved water security (quantity, quality and access) in 12 households in a Guatemalan peri-urban village and the potential to expand this approach regionally and beyond. Glasgow et al. [46] assessed respondents’ knowledge, attitudes and practices using household rainwater harvesting systems in 20 homes in Carricoao in Grenada.

Both studies reported increased water quantity. Respondents in Elgert’s study noted using rainwater for almost six months of the year, meaning family members (primarily women) reduced the number of trips made to the public water points compared to those without the rainwater harvesting system. Half of the participants in Glasgow et al.’s study reported water availability throughout the year. In Guatemala, rainwater was primarily used for drinking and cooking, whilst laundry was done at public water points to conserve rainwater [53]. In Carricoao, respondents used rainwater for drinking, cooking, bathing, laundry and cleaning [46]. Individual household water treatment practices were not gathered in Elgert et al.’s study, but the authors recommended boiling water before consumption. Respondents in Glasgow et al.’s sample reported that just over half treated water before drinking by adding bleach chlorine tablets and using small fish to consume insects. No data on water-related or borne diseases was gathered in either study.

Both studies concluded that rainwater harvesting systems could increase water security but that regular operation and maintenance of the system is critical; additionally, Glasgow et al. highlighted the importance of educational campaigns to improve water treatment practices before consumption.

### Role of the government and service providers

Nine papers mentioned government and service providers (non-government organisations and urban utilities) [38–40, 43, 47, 51, 53, 56, 60]. No papers included data about water user groups or committees. Nayebare et al. [43] set out the government’s roles and responsibilities but provide no other information. Five of these papers detail the technology the government and service providers install or subside to increase water quantity and quality and, therefore, resilience to weather events [38, 39, 53, 56, 60]. These include household rainwater harvesting in Guatemala [53], chlorinated piped water in Dar es Salaam [56], pond sand filtration, reverse osmosis, household rainwater harvesting, deep tubewells with overhead water storage, and the provision drinking water in 10-40 litre containers in Bangladesh [38, 39, 60]. Ahmed et al. [40] report perceptions of government mismanagement of the water supply. Ashrafuzzaman and Furini [38] indicated that governments and NGOs rarely delivered pond sand filtration in collaboration with community members in remote rural areas. Iyer and Toe [51] highlight that funding from the local government for sanitation is insufficient, leading to the construction of makeshift latrines that are easily damaged in weather events. Nor do they provide funding for retrofitting damaged latrines.

## Discussion

We aimed to explore how climate change impacts WASH services and behaviours and if climate-resilient WASH interventions are disability-inclusive and gender-equitable in LMICs. The findings underscore how vulnerable WASH services and behaviours are to climate change. Studies included evidence about how the changing climate impacts WASH services and behaviours. However, we identified a significant gap in epidemiological research linking these impacts to specific health outcomes. Furthermore, we found a dearth of evidence related to the experiences of people with disabilities. More data was found regarding women and girls, and some considered how climate impacts gender relations. Notably, the review identified a scarcity of evidence regarding the effectiveness of climate-resilient WASH interventions at the household and community levels. Only two studies explored rainwater harvesting systems, highlighting the need for further research on effective adaptation strategies.

No previous reviews have been conducted on this topic, yet we found only 22 papers that met our inclusion criteria, demonstrating that this is an emerging work area. Rainfall uncertainty, drought, increased rainfall and flooding were the most commonly documented weather events in the studies. Though most papers examined the effects on various aspects of water access, the split across water, sanitation and hygiene was marginal. This demonstrates the recognition that, holistically, water, sanitation, and hygiene services and behaviours contribute to maintaining public health during weather events.

Although the impacts on women and girls were present in over half the studies, evidence related to people with disabilities remains scarce. This pattern is observed in studies that assessed disability inclusion in WASH-related policies in Nepal, Bangladesh, and Cambodia compared to gender [61, 62]. Across all countries, attention to disability was considerably less than to gender. Both populations are vulnerable to the risks of climate change, so studies relating to these populations must be equally increased. As identities intersect to deepen poverty and marginalisation, attention must also be given to women and girls with disabilities in future studies.

Furthermore, where disability is mentioned, the population was referred to as a homogenous group without reference to the impairment experienced or gender [49], or the emphasis was on physically accessing the WASH service for persons with sensory or physical disabilities [58]. A steadily growing body of evidence has marked the importance of understanding how intersecting identities shape WASH experiences [11, 63, 64]. These report the nuanced barriers faced by women and men with disabilities of different ages across the different impairment groups when accessing WASH services, which makes designing appropriate solutions more feasible [8]. This must now include climate change WASH service adaptation and resilience efforts.

Our review paints a concerning picture of the impact of extreme weather events on water usage, sanitation and hygiene behaviours, and health outcomes. Across all weather events, many people resorted to consuming untreated water, diversifying water sources and using them for different purposes (e.g. rain or pond water for cooking and personal hygiene). These are common coping strategies in many LMIC settings [6, 65, 66]. It is recognised that having access to diverse water sources increases climate resilience. However, the focus must be on having access to diverse water sources of good quality and close proximity [67].

Our review also highlights the importance of social capital and kinship networks during water scarcity. However, people with disabilities are particularly vulnerable during droughts, as they often rely on social networks’ moral obligation to collect water. Arguably, this could decline as the situation worsens for everyone. Such vulnerability to more significant inequalities underscores the need for targeted support mechanisms to ensure access to water for persons with disabilities during droughts.

Kinship and social capital have always been important in distributing finite resources, but the latter has been identified as a potential strategy to build resilience to climate change [68, 69]. While social capital is crucial for immediate survival, further research is needed to understand its broader effects on climate change adaptation. More effective strategies to support communities during droughts can be designed by understanding these coping mechanisms and their limitations. These include promoting sustainable water management practices, bolstering social support networks, and ensuring targeted assistance for populations that are vulnerable to exclusion.

A key finding from this review is that households prioritised water for agriculture and livelihood security over good sanitation and hygiene practices. This prioritisation leads to increased rates of open defecation in most settings and a decline in hygiene as households limit water use for activities like handwashing, utensil cleaning, laundry, and menstrual health and hygiene. Critically, we found no data on the menstrual health experiences of women and girls with disabilities. While growing evidence underscores the importance of menstrual health for all women and girls, including in humanitarian crises, the intersection of menstrual health, disability and climate change is unexplored [70–75]. To effectively address menstrual health challenges exacerbated by climate change, it is imperative to consider the unique requirements of women and girls with disabilities and caregivers who support them. Failure to do so will only deepen existing menstrual health inequalities.

Increased rates of illness reported across different settings include diarrhoea, abdominal pain, headache, dehydration, intestinal worms (rainfall uncertainty and drought, increased rainfall, flooding, and cyclones), high blood pressure, hypertension, respiratory infection, pre-eclampsia, rashes, and urinary tract infections (climate change-induced sea level rise and saltwater intrusion). Diarrhoea disease is the second leading cause of death and the leading cause of malnutrition in children under five years old [76]. There are significant disparities in severe diarrhoea rates, with children under five with disabilities experiencing twice the rate as those without disabilities in the same age group [9, 77]. However, none of the reviewed papers reporting increased diarrhoea rates considered disability—a major oversight given the health inequalities faced by people with disabilities. Diarrhoea is treatable and preventable. A recent systematic review found that community-wide sanitation prevents the spread of diarrhoea [5], and handwashing with soap reduces diarrhoeal disease by 30% [2, 4]. With human-induced climate change causing more intense and extreme weather events [31], it is now more vital than ever to invest climate finance into increasing access to WASH services that are resilient to weather events and developing and maintaining positive WASH behaviours.

Declining mental health and wellbeing also came out in our results. This chimes with the expanding evidence on climate change and mental health: a scoping review by Charlson et al. [78] reported that climate-related exposures, including increased rainfall, drought and floods, were linked to poor mental health outcomes, including increased distress, hospitalisations, and suicide rates, especially for those with pre-existing mental health conditions. Wahid et al. [79] explored the association between climate-related shocks and depression in Bangladesh. The authors found that exposure to floods in the last year was associated with depression and anxiety, and the odds were significantly higher for people with physical disabilities than those without disabilities. Our review did not uncover any further data related to disability and mental health, so more research is required to understand the intersection between disability, climate change, and mental health.

Much evidence exists about the connections between safely managed WASH and gender equality. Women and girls are more likely to collect water, which impacts time spent in school or at work [7]. Collecting water is associated with stress, fatigue, pain, perinatal health problems and vulnerability to violence, including sextortion [80–84]. Our review shows how changing weather events will likely exacerbate these issues. Furthermore, papers in our review indicate that water scarcity negatively impacted lactating mothers, leading to dehydration and decreased milk production. Though evidence exists, Miller et al. [85] note that a more rigorous exploration of the issues is required.

In our review, climate change adaptation activities aimed at reducing vulnerability to the present and future impacts of climate change include pond sand filtration, Reverse Osmosis, buying water and installing piped water, protecting homesteads with flood defences, constructing latrines on higher ground and constructing latrines with more durable materials. Yet, affordability was a major barrier for many people, highlighting the need for financial and social safety nets to ensure equitable access to these critical WASH interventions. Such efforts must specifically target persons with disabilities because this population often experiences greater poverty than those without disabilities. A recent study using the global Multidimensional Poverty Index found that in 45% of the countries studied, households with persons with disabilities face higher levels of poverty than those without [86].

Only two papers included assessments of climate-resilient WASH services, and both focused on rainwater harvesting, a technique that has been applied for generations. Though both studies demonstrate its potential to increase water quantity, water treatment was limited, and neither study examines waterborne disease prevalence or developing and maintaining WASH behaviours, which presents an evidence gap. Evaluations of climate-resilient WASH services should comprehensively assess the technology employed, user behaviours, the effectiveness of system maintenance practices, and historical rainfall data incorporating long-term trends [87].

A recent study investigating the impact of seasonal rain patterns and extreme weather events on household water security in rural Gambia highlighted potentially climate-resilient WASH infrastructure, including solar-powered boreholes [66]. Though users reported less vulnerability to water sources, households faced shortages during the rainy season with greater cloud coverage. The complex interplay between mechanical failures, weather events and agricultural practices contributed to water insecurity.

However, evaluating WASH technologies will not suffice; we must understand the effectiveness of increasing and sustaining positive WASH behaviours and develop transformative approaches that tackle deeper social and economic issues to achieve climate justice and inclusion. A key aspect of climate-resilient WASH is a population’s ability to absorb and recover from weather events, making the efforts of governments, service providers, and communities to increase resilience vital. Unfortunately, minimal data exists related to these efforts. We hypothesize that activities are occurring but have not yet been evaluated and published. Therefore, scientific evaluations of interventions aimed at increasing climate-resilient WASH are urgently required to determine what components successfully enhance the resilience of persons with disabilities to weather events that impact their WASH experiences. This proposal aligns with a recommendation from a recently published review on disability and WASH, which outlines a roadmap for progressively realising the rights of persons with disabilities to WASH by 2030 [8](page 8).

### Strengths and limitations of review

Our search for studies on WASH and disability was thorough, but limitations may exist. Qualitative studies, especially those without explicitly mentioning disability in the title or abstract, could have been missed. Additionally, the relatively new field of climate-resilient WASH might have resulted in overlooking relevant evaluations due to a lack of standardised terminology. This highlights the need for a clearer definition of climate-resilient WASH to facilitate future research and ensure all pertinent studies are captured.

## Conclusion

This review underscores the critical intersection between climate change and WASH services, highlighting the disproportionate impact on vulnerable populations, especially people with disabilities, women, and girls. Our findings reveal a significant gap in understanding the health consequences of climate-related WASH disruptions, particularly for people with disabilities. The review identifies a pressing need for more comprehensive studies on effective climate-resilient WASH interventions at the household and community levels. The prioritisation of water for agriculture over sanitation and hygiene, coupled with the coping mechanisms employed during extreme weather events, underscores the complex challenges communities face. The review underscores the need for targeted support mechanisms, sustainable water management practices, and financial and social safety nets to ensure equitable access to WASH services. Substantial efforts are needed to expand the evidence base. This includes rigorous evaluations of climate-resilient WASH technologies and transformative approaches that address underlying social and economic issues. By prioritising inclusive and effective WASH interventions, we can enhance climate resilience and promote health equity for all, particularly the most vulnerable populations.

## Supporting information

S1. PRISMA Scoping Review Checklist

S2. Search strategy and key terms

## Data Availability

All data produced in the present work are contained in the manuscript.

## Acknowledgements

Thank you to Sabiha Ahmed and Mahbub Ul Alam for contributing to the broader research exploring how human-induced climate change impacts people with disabilities’ WASH experiences in Bangladesh.

## Authors contribution

Conceptualization: Jane Wilbur, Julian Natukunda

Data curation: Jane Wilbur, Julian Natukunda

Funding acquisition: Jane Wilbur

Investigation: Julian Natukunda, Jane Wilbur

Methodology: Julian Natukunda, Jane Wilbur

Project administration: Jane Wilbur

Supervision: Jane Wilbur

Writing – original draft preparation: Jane Wilbur

Writing – review & editing: Jane Wilbur, Julian Natukunda, Doug Ruuska, Shahpara Nawaz

