## Supplementary material for "Climate Risks to Water, Sanitation and Hygiene Services and Evidence of Inclusive and Effective Interventions in Low and Middle-Income Countries: A Scoping Review": S2. Search strategy and key terms

| Weather events | MeSH terms: WeatherClimate change, Disasters  (rain* OR windstorm OR storm* OR hurricane* OR "extreme weather" OR typhoon* OR cyclone* OR "sea level" OR "coastal erosion" OR snow OR "storm surge" OR "high tide" OR "pluvial flood*" OR "heavy rainfall" OR “extreme rainfall” OR “low rainfall” OR "fluvial flood*" OR lightning OR monsoon OR drought OR "saline flooding" OR "sea level rise" OR "river flood*" OR "surface water flood*" OR "surface flood*" OR "coastal flood*" OR "flood* event*" OR "natural disaster" OR “climate change”) |
| --- | --- |
| WASH terms | “water service”, “water access”, “water supply”, “water suppl*”, “drinking water”, “household water”, “domestic water”, “sanitation”, “open and defecation”, “wastewater”, “waste and water”, “sewer*”, “hygiene”, “WASH”, “menstrual and health”, “menstrua*”, “menstrual and hygiene and management”, “menstrual and hygiene”, “menstrual and health and hygiene”, “handwashing”, “hand AND washing” |
| Evaluation terms | “evaluation AND trial”, “effectiveness AND trial”, “outcome AND evaluation”, “impact AND evaluation”, “impact AND study”, “feasibility AND study”, “feasibility AND acceptability AND study” “longitudinal AND study”, “mid AND term AND review”, “program* AND evaluation”, “program* AND review”, “program* AND trial”, “summative AND assessment”, “clinical AND review”, “clinical AND trial” |
| Disability | Disabled person OR disabled person* OR person with disabilit* OR persons with disabilit* OR people with disabilit* OR handicapped person* OR handicapped people, Physical impair* or physically impair*, OR Deaf* or hearing loss OR hearing impair*, Blindness OR vision loss* OR vision impair*, psychosocial OR cogniti* and disabilit*, OR cogniti* and impair*, dementia* or Alzheimer*, intellectual and disabilit* OR intellectual impair*, OR mentally ill OR mental illness*OR mental impair*, OR developmental disabili*, Learning disorders OR learning disorder* OR communication disorder*, autistic OR autism OR asperger* OR Down’s Syndrome OR Down Syndrome |

**Grey literature key words**

| **Keywords** | **synonyms** |
| --- | --- |
| A.Weather events | “extreme weather” or “climate change” or “natural disasters |
| B. Water, sanitation, and hygiene | WASH or “Menstrual health and hygiene” or “climate-resilient WASH interventions” |
| C. effectiveness | Feasibility or evaluation |
| D. People with disabilities | “People living with disabilities” or “mental illness” or “mental disorders” |
